# Respiratory symptoms and behavior change in people with Down syndrome during the social lockdown

**DOI:** 10.1101/2025.11.20.25340480

**Authors:** E. de Vries, N.B. Eijsvoogel, B.M.M. Streng, A.M. de Groot, L.J. Bont, J.G. Wildenbeest, G.Th.J. van Well

## Abstract

During the COVID-19 pandemic and associated social lockdown, a reduction in positive respiratory viral testing was noted in the general population, with increased depression and loneliness and decreased life quality. Respiratory symptoms occur more frequently in Down syndrome (DS) with multifactorial causes. The aim of this study was to analyze the reduction in respiratory symptoms in DS during the pandemic to increase our understanding about the proportion of respiratory symptoms attributable to infections in DS. The degree of reduction in respiratory symptoms could also have influenced behavior and well-being in DS. We studied this in the PRIDE cohort. The decrease in respiratory symptom frequency was inversely related to age in DS. We observed mixed changes in behavior and/or well-being in about half the DS participants (the changes were not always negative) and they were not clearly related to respiratory symptom frequency. In conclusion, our results suggest that respiratory symptoms in DS are more often related to viral infections in DS children, but less often in DS adults.

## INTRODUCTION

Respiratory symptoms occur more frequently in Down syndrome (DS) [1]. Contributing factors are an abnormal immune system, abnormal physiology (e.g., abnormal function of cilia, hypotonia) and anatomical differences (e.g., midface hypoplasia, tracheolaryngomalacia) [e.g., 1]. The influence of respiratory infections on quality of life can be considerable in Down syndrome children [2]. The course of respiratory infections is often more severe in DS with higher morbidity and mortality [3].

During the COVID-19 pandemic and associated social lockdown, a general reduction in positive respiratory viral testing was noted and the yearly RS and influenza virus winter peak was absent [4]. The downside of the social lockdown was the increase in depression and loneliness and the decrease in quality of life (QoL) [5]. In people with DS, behavioral problems, mood swings, depression and a lower QoL were also observed [6].

The aim of this study was to analyze the reduction in respiratory symptoms in DS during the pandemic to increase our understanding about the proportion of respiratory symptoms attributable to infections in DS. Given the multifactorial cause of respiratory symptoms in DS (i.e., not only based on infections), it could be that their reduction in respiratory symptoms had been lower during the social lockdown than in the general population. Next to the general downside of the lockdown, this could also have influenced their behavior and well-being. We were able to study this in the large cohort of the PRIDE project (Prospective Monitoring of Antibody Response Following COVID-19 Vaccination in Patients With Down Syndrome) [7].

## METHODS

People with DS and their household contacts and healthcare workers (as controls) took part in the PRIDE project after written informed consent (approved by METC UMCU; NL76336.041.21). As a side study, participants were later asked to complete an online survey about their respiratory symptoms and changes in behavior and general well-being during the social lockdown.

Survey questions were composed in collaboration between all authors based on the available relevant literature, and where necessary adapted to the age category of the participants.

The survey questions were incorporated in Castor EDC (8) and the survey fitting the age category of each potential participant was sent by email. The email contained a link to the online questions, and the answers were directly entered into the Castor system. A reminder email was sent after two weeks. All available data were exported and analyzed. Quantitative data were analyzed using descriptive statistics. Qualitative data (the final open text question) were analyzed by the first author, using inductive coding and thematic grouping thereafter [9].

## RESULTS

### Participants

The survey invitation email was sent to the 347 PRIDE participants and was fully completed for 126 DS (5-11y n=13, 12-19y n=12, 20-39y, n=84, ≥40y n=17) and by 58 control participants (full response rate 51%; 15 (4%) partially completed questionnaires were excluded; participant characteristics in Table 1; overview of daily activities in Figure 1.

**Table 1.** Baseline characteristics of the participants.

|  | DS 5-11y | DS 12-19y | DS 20-39y | DS ≥40y | controls |
| --- | --- | --- | --- | --- | --- |
| Asthma in family (n, %) | 4<sup>1</sup> (31) | 2 (17) | 21<sup>2</sup> (25) | 4<sup>2</sup> (24) | 10 (20) |
| Eczema in family (n, %) | 5 (38) | 6 (50) | 36<sup>2</sup> (43) | 5<sup>2</sup> (29) | 9<sup>1</sup> (18) |
| Allergy in family (n, %) | 7 (54) | 9<sup>1</sup> (75) | 42<sup>3</sup> (50) | 6<sup>3</sup> (35) | 22 (43) |
| Smoking in household (n, %) | 1 (8) | 0 (0) | 2 (2) | 0<sup>1</sup> (0) | 0 (0) |
| Congenital heart defect (n, %) | 6 (46) | 5 (42) | 35<sup>2</sup> (42) | 6<sup>1</sup> (35) | 0 (0) |
| Hypotonia (n, %) | 7<sup>1</sup> (54) | 8<sup>2</sup> (67) | 28<sup>4</sup> (33) | 1<sup>5</sup> (59) | 0 (0) |
| Laryngomalacia (n, %) | 1<sup>6</sup> (8) | 2<sup>6</sup> (15) | 3<sup>7</sup> (23) | 1<sup>5</sup> (8) | 0 (0) |
| Sex (f; n, %) | 3 (23) | 9 (75) | 38<sup>8</sup> (56) | 0 (0) | 41 (71) |

<sup>1</sup>1 unknown; <sup>2</sup>2 unknown; <sup>3</sup>3 unknown; <sup>4</sup>6 unknown; <sup>5</sup>4 unknown; <sup>6</sup>3 unknown; <sup>7</sup>14 unknown; <sup>8</sup>16 unknown%, percentage; DS, Down syndrome; f, female; n, number; y, years-of-age

**Table 2.** Respiratory symptom frequency changes during the lockdown.

| group | n | symptom frequency change |  |  |  |
| --- | --- | --- | --- | --- | --- |
|  |  | no change | fewer | more | missing |
| 5-11y DS | 13 | 4 (31) | 9 (69) | 0 (0) | 0 (0) |
| 12-19y DS | 12 | 7 (58) | 5 (42) | 0 (0) | 0 (0) |
| 20-39y DS | 84 | 54 (65) | 23 (27) | 5 (6) | 2 (2) |
| ≥40y DS | 17 | 14 (82) | 1 (6) | 2 (12) | 0 (0) |
| adult controls | 51 | 34 (67) | 13 (25) | 1 (2) | 3 (6) |

DS, Down syndrome; n, number; y, years-of-age. Symptoms: number (percentage).
Answer to the question *How many respiratory complaints did he/she / did you have during that strict lockdown? Rules during the strict lockdown from January 2021 to May 2021: complete lockdown; curfew; all schools closed; homeschooling; closure of childcare facilities; closure of non-essential shops.*

**Figure 1.**
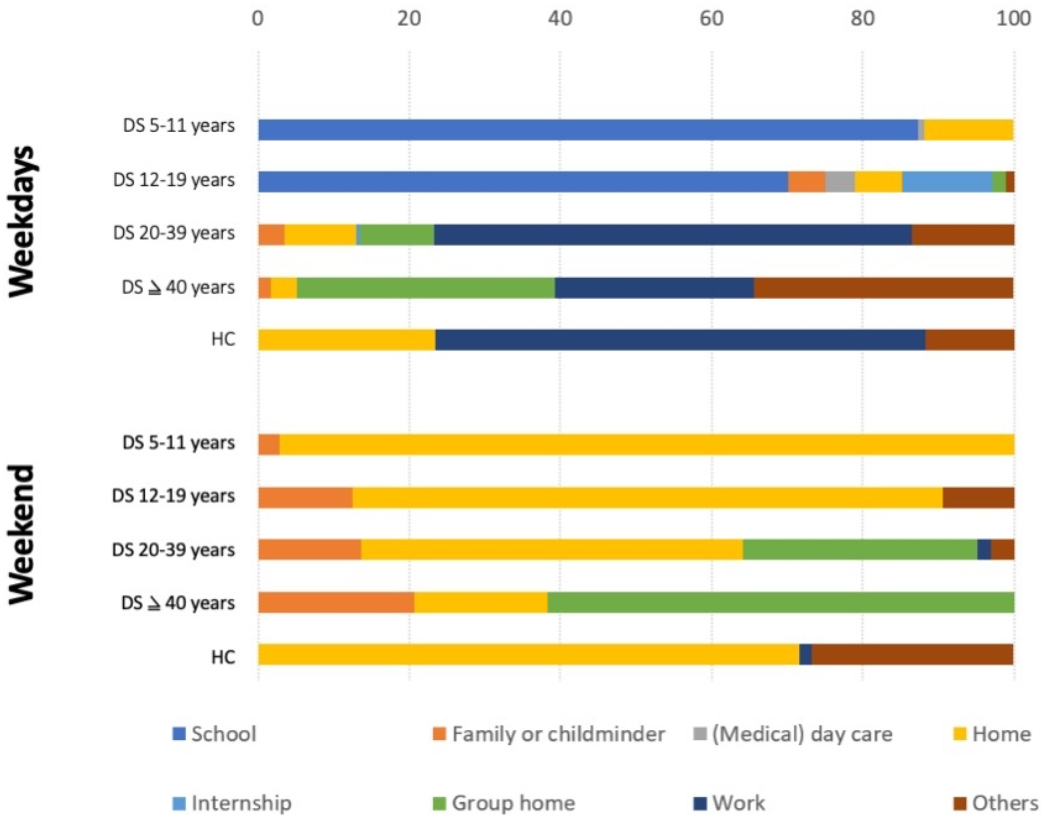
Overview of daily activities of the participants (outside of the pandemic period).

### Respiratory symptoms

Respiratory symptoms were generally decreased during the pandemic. The decrease was inversely correlated with age and in adults above 40 years this effect was no longer present.

### Behavior and well-being

In around half of the participants, behavior and/or well-being changes during the lockdown were reported (see Table 3) and described in the free text box (analysis of the results see below).

**Table 3.** Behavior and well-being changes during the lockdown.

| group | n | behavior and well-being change |  |  |  |
| --- | --- | --- | --- | --- | --- |
|  |  | yes | no | don't know | missing |
| 5-11y DS | 13 | 6 (46) | 6 (46) | 1 (8) | 0 (0) |
| 12-19y DS | 12 | 6 (50) | 6 (50) | 0 (0) | 0 (0) |
| 20-39y DS | 84 | 36 (43) | 45 (54) | 1 (1) | 2 (2) |
| ≥40y DS | 17 | 8 (47) | 7 (41) | 2 (12) | 0 (0) |
| adult controls | 51 | 20 (39) | 23 (45) | 5 (10) | 3 (6) |

DS, Down syndrome; n, number; years-of age. Symptoms: number (percentage).
Answer to the question *Has the lockdown and staying at home affected your [controls] behavior and general well-being / the behavior and general well-being of the person with Down syndrome?*

#### Mood and behavior

A good, or even better, mood during the lockdown due to the less-demanding period within the family home or sheltered living facility was described for around one third of DS children/teenagers, but only for around 10% of DS adults (and 15% of controls).

> “After a period of getting used to the disruption of his daily routine, being home a lot with the whole family actually gave him a lot of peace. He was his cheerful self.” (parent of 5-11 year-old DS child) “He thought it wsas fine. The peace, the regularity. No ‘musts’.” (parent of 20-39 year-old DS adult)

In around a quarter of DS children and adults, missing school, friends, sports, day care and/or work negatively influenced their mood and destabilized them, leading to stubborn behavior, restlessness, anger, and aggression (also described in 14% of controls). DS participants were increasingly described as being bored, gloomy, anxious, listless, lonely, and/or withdrawn with increasing age.

> “He didn’t get it, especially about wearing the face mask. That took way too long, was unclear. It made him restless and sometimes aggressive. (parent of 20-39 year-old DS adult)
>
> “Very stressed, restless and sometimes with mild aggression due to extreme frustration.” (adult control)

#### Performance and development

One half of school age DS children showed a decreased learning performance in the lockdown. Social and emotional development was negatively influenced in around 13% from school age onwards.

> “I think it had an impact that I can’t yet fathom, …, social behavior has been adjusted, development is different than from that moment on, etc.” (parent of a 12-19 year-old DS teenager)
>
> “The consequence of the lockdown is that he was very bad at reintegrating afterwards and no longer wants to participate in many activities, this is still the case, socially he has lost a lot of it.” (parent of 20-39 year-old DS adult)

#### Physical fitness

Physical fitness improved during the lockdown in 17% of DS children/teenagers but deteriorated in 50% of them. DS adults did not improve in their physical fitness; it deteriorated in 9% of them, also leading to increased body weight.

> “She preferred to hang out on the couch all the time.” (parent of 12-19 year-old DS teenager) “… weight gain due to not being able to exercise daily.” (parent of 20-39 year-old DS adult)

## DISCUSSION

We were able to investigate the frequency of respiratory symptoms in a Down syndrome cohort and healthy controls from the PRIDE study using a survey sent by email to the participants of this study. Our primary objective was to determine the effect of the social lockdown during the COVID-19 pandemic on the frequency of respiratory symptoms in the participants of the PRIDE study.

Respiratory symptoms were generally decreased during the pandemic. The decrease was inversely correlated with age and in adults above 40 years this effect was no longer present. These results suggest that respiratory symptoms in DS are less often caused by viral infections with increasing age.

Unfortunately, the response rate to the survey was lower than expected. Therefore, it was not possible to perform an analysis of each separate respiratory symptom.

Changes in behavior and/or well-being were described in about half the DS participants, but these were not always negative changes. This finding highlights the heterogeneous composition of the DS community.

In conclusion, our results suggest that respiratory symptoms in DS are more often related to viral infections in DS children, but less often in DS adults.

## Data Availability

All data produced in the present study are available upon reasonable request to the authors.

https://www.tilburguniversity.edu/nl/medewerkers/e-devries

## Notes

### Competing Interest Statement

The authors have declared no competing interest.

### Funding Statement

This study was supported by ZonMw, The Netherlands Organization for Health Research and Development, grant 10430072010004.

### Author Declarations

The Ethics committee of University Medical Center Utrecht evaluated the survey study and gave a waiver from the "Wet Medisch Onderzoek bij mensen" (the Dutch law regulating ethical evaluation of medical research in humans).

